# Phase-targeted modulation of essential tremor with transcranial magnetic stimulation of motor cortex

**DOI:** 10.64898/2026.05.11.26347791

**Authors:** Valentina Mancini, Isaac Grennan, Nicolas Shackle, Talia Vasaturo-Kolodner, Priya Sharma, Alina Siekmann, Sagel Kundieko, Fabio Ferrandes, Lara Biller, Karen Wendt, Kawsar Ali, Dan Rogers, Nagaraja Sarangmat, Ashwini Oswal, Timothy Denison, Hayriye Cagnan, Andrew Sharott, Charlotte J Stagg

## Abstract

Neural oscillations provide temporal frameworks for coordinating communication within and across distributed brain networks. In essential tremor (ET), pathological synchronization within the cerebello–thalamo–cortical circuit produces rhythmic activity that manifests as an involuntary action tremor. Although deep brain stimulation can effectively suppress tremor, its invasiveness and cost highlight the need for non-invasive interventions capable of selectively modulating pathological oscillations.

Transcranial magnetic stimulation (TMS) offers a non-invasive means of engaging cortical circuits, yet conventional stimulation protocols are delivered independently of the ongoing neural dynamics. Such open-loop approaches ignore the temporal structure of tremor-related activity, potentially stimulating during both amplifying and suppressing phases of the oscillation. To address this, we compared two phase-targeted TMS paradigms: first-pulse phase-locked TMS (First-pulse -TMS), in which only the initial pulse of a stimulation train is aligned to the tremor phase, and cycle-by-cycle phase-locked TMS (Continuous-TMS), in which each pulse is continuously triggered based on real-time tremor phase.

Ten patients with ET underwent stimulation guided by peripheral tremor recordings using an accelerometer, with tremor phase estimated in real time via the Oscilltrack algorithm. Sixty-four trains of TMS pulses were delivered at nine discrete phase bins of the tremor cycle, such that each phase bin was repeated approximately seven times. Continuous-TMS maintained accurate phase-locking across consecutive cycles (mean phase-locking value ∼0.9), whereas First-pulse-TMS exhibited progressive drift over time and low phase consistency (mean phase-locking value <0.2). The circular concentration of stimulation phase was significantly greater for Continuous-TMS than First-pulse-TMS (Mann–Whitney U-test, p < 0.001), indicating a significant difference in overall phase-locking accuracy between the two protocols. Critically, Continuous-TMS, unlike First-pulse-TMS, induced bidirectional, phase-dependent modulation of tremor amplitude. Circular-linear modelling revealed a sinusoidal relationship between stimulation phase and changes in tremor amplitude, with tremor amplification and suppression occurring at opposite phases of the cycle. Covariates including baseline tremor amplitude and trial number were accounted for. In some people, tremor suppression outlasted the stimulation period, suggesting phase-locked TMS may be a potentially useful therapeutic tool.

By enabling reliable, phase-specific stimulation of the tremor cycle, Continuous-TMS allows identification of the individual phase that produces maximal tremor suppression, supporting the development of personalized, phase-specific neuromodulation strategies. This proof-of-principle study demonstrates that temporally precise, closed-loop TMS can interact with pathological oscillations in real time, providing a mechanistic framework for probing oscillatory contributions to motor symptoms and a scalable therapeutic approach for ET and other oscillopathies.

## Introduction

Essential tremor (ET) affects approximately 1% of the adult population globally, classically presenting with an action tremor in one or both upper limbs. ET is often disabling, and pharmacological treatments to alleviate symptoms are only partly effective, with up to one third of ET patients deciding to stop pharmacological treatment. Conversely, the placement of a thalamic deep brain stimulation (DBS) electrode is often highly effective at reducing tremor and hence restoring quality of life but is a highly limited and resource-intensive approach.

ET is a prototypic oscillatory disorder, arising from pathological entrainment of neural populations within the cerebello-thalamo-cortical network, which results in abnormally enhanced oscillatory activity that interferes with normal motor output. Oscillations are ubiquitous in neural systems, providing frequency-dependent temporal reference frames that are optimal for different modes of communication and coordination within and across brain areas. Oscillations in different frequency bands appear to play a variety of broad computational roles that are maintained across different cognitive domains^1-3^. However, in many brain diseases, the power and synchronisation of specific oscillatory activities is altered, and these changes can provide valuable readouts of disease processes and symptoms^4,5^.

Non-invasive brain stimulation (NIBS) approaches that can precisely modulate oscillatory activity may provide a potentially powerful method of manipulating pathological processes such as tremor. In particular, phase-targeted, closed-loop methods, whereby real-time processing of the signal is used to control the timing of stimulation^6^ provide particularly powerful approaches, because they can amplify or suppress ongoing oscillations depending on the precise phase of the underlying oscillation being targeted^7-9^. This in turn can modulate the motor and cognitive dysfunctions associated with those oscillations. For example, phase-dependent modulation of parkinsonian beta oscillations in dopamine-depleted rats leads to altered locomotor speed and pattern^10^. Similarly, phase-targeted modulation of theta oscillations in memory circuits can powerfully modulate cognitive performance^11-15^. We have recently demonstrated that optogenetic phase-dependent stimulation of motor cortical neurons can also modulate the amplitude of motor cortical theta oscillations^16^, providing proof-of-concept evidence that such an approach could work at the level of cortical circuits.

Although the potential advantages of NIBS over implanted devices are clear, the effects of non-invasive stimulation are weakened by inevitable loss of energy reaching the brain. Therefore, increasing effect size by delivering stimulation at specific phases of relevant ongoing oscillations is a highly promising approach to improve their efficacy. This is particularly true for transcranial magnetic stimulation (TMS), which provides more temporally discrete pulses than other non-invasive brain stimulation approaches. However, TMS leads to significant artefacts in the EEG signals that provide the most readily available input to closed-loop systems, preventing accurate phase-tracking of oscillations above 2Hz.

TMS-induced artifacts in EEG signals have posed a significant barrier to the implementation of continuous closed-loop brain stimulation, limiting the availability of real-time feedback throughout stimulation periods. To circumvent the disruption of phase estimation by these artifacts, prior studies have employed strategies wherein isolated TMS pulses or pulse trains are triggered based on the phase of EEG oscillations detected during artifact-free periods when the stimulation is off^17,18^. A first-pulse phase-locked TMS (First-pulse-TMS) has previously been demonstrated^19^. With this approach, the first pulse of TMS is phase-locked to the alpha-oscillation in the EEG but all subsequent pulses of TMS are delivered in an open-loop fashion at a constant inter-stimulation interval matched to the period of the alpha oscillation. This aims to continue to maintain phase-locking across the TMS train, without real-time phase-tracking. However, this lack of closed-loop feedback during First-pulse-TMS almost inevitably means that phase precision degrades across the stimulation train.

Overcoming these limitations using peripheral signals would allow proof-of-principle investigations as to whether cycle-by-cycle phase-locked TMS (Continuous-TMS) is a better approach to manipulate neural oscillations. Continuous-TMS is a highly tractable approach for ET as tremor phase is highly synchronised with the phase of ongoing pathological oscillations in the cortico-thalamo-cerebellar circuit from which it originates^20^. Stimulating on a given phase of tremor thus acts as a proxy for the neural signals that generate tremor but without the large TMS-artefacts present in EEG. Indeed, using this approach ET and Parkinsonian tremor can be suppressed using phase-targeted transcranial alternating current stimulation (tACS)^7^ and thalamic deep brain stimulation (DBS)^21^.

Here we examined for the first time whether the amplitude of tremor in ET patients could be modulated using continuous-TMS. To achieve this, we used our recently developed phase-tracking algorithm (Oscilltrack^22^) to estimate the phase of peripherally recorded tremor oscillations in real-time and used the output for an existing First-pulse-TMS paradigm^19^ and novel continuous-TMS using the next generation xTMS device. Our results demonstrate that continuous-TMS can be accurately targeted to specific phases of ongoing tremor oscillations, leading to amplitude modulation that is dependent on cycle-by-cycle closed-loop effects. Further, in a subset of patients both First-pulse-TMS and continuous-TMS led to sustained tremor suppression. This proof-of-principle implementation highlights the potential of continuous-TMS to enhance the clinical efficacy of closed-loop stimulation of cortical areas, which could provide novel therapeutic strategies for a range of brain disorders.

## Methods

### Participants

This study was reviewed and approved by the National Research Ethics Service (North West Liverpool Central REC reference: 20/NW/0150), adhering to the guidelines set forth in the Declaration of Helsinki, except for preregistration. All participants provided their written informed consent to take part in the study,

Twelve participants diagnosed with essential tremor (ET) were recruited through the Oxford University Hospitals NHS Trust or via the National Tremor Foundation. We excluded patients with diagnoses of other neurological conditions and contraindications to transcranial magnetic stimulation (TMS) such as a history of seizures or the presence of metallic implants above the waist, cardiac pacemakers or insulin pumps.

Two participants (S08 and S12) had an insufficiently strong distal tremor amplitude in their most effected upper limb (defined as less than 0.2 m/s^2^ according to the Bain and Findley tremor severity scale ^23^) and were excluded from further analysis. The final study cohort therefore consisted of ten participants with ET (four female; mean age = 62.8 ± 15.9 years; age range = 25 – 84 years; mean time since diagnosis = 13.3 ± 9.2 years (see Table 1 for detailed demographic information).

### Study outline

We employed a within-participants cross-over design: each participant attended two sessions: one using First-pulse-TMS and one with continuous-TMS, the order of which was counterbalanced across the group. Two participants were able to attend only one session. Participants were asked to abstain from tremor-related medications on the morning of the experiment.

In each session, data were recorded from the upper limb that exhibited the most pronounced tremor, and participants were asked to assume a tremor provoking position during TMS blocks. Tremor amplitude was recorded: (i) at baseline in the absence of stimulation, (ii) during 16 blocks of TMS, and (iii) after stimulation. A triaxial accelerometer (Biometrics Ltd, ACL300) and electromyographic (EMG) surface electrodes were used to monitor tremor dynamics and muscle activity throughout the experiment. EMG electrodes were placed on the first dorsal interosseous (FDI), abductor pollicis brevis (APB), forearm finger flexors (FFF), and forearm finger extensors (FFE). EMG activity was sampled at 5 kHz, amplified using a Digitimer D440 4-channel Amplifier (Digitimer Ltd.), and processed with CED Power1401 Signal Software (Cambridge Electronic Design, UK). The accelerometer was positioned between the first and second metacarpal heads on the dorsum of the hand, with the signal sampled at 1 kHz and processed using the same data acquisition software as the EMG data.

### TMS procedure

TMS was applied to the motor hotspot for the First Dorsal Interosseous (FDI) within the primary motor cortex (M1) contralateral to the hand with the strongest tremor, with the TMS coil oriented held posteriorly 45 degrees to the midsagittal line. The resting motor threshold (rMT) and active motor threshold (aMT) were defined for each participant in each session. The rMT was defined as the lowest intensity required to elicit a peak-to-peak MEP of at least 50 μV in the FDI for 5 out of 10 single pulses. The aMT was defined as the minimum stimulus intensity needed to produce an MEP with a peak-to-peak amplitude of at least 200 μV in 5 out of 10 trials, during isometric contraction of the tested muscle at approximately 20% of maximum voluntary contraction^24^. All TMS pulses were delivered at 90% aMT.

A maximum of 960 pulses were delivered per session, and no participants reported any adverse effects either during or after TMS with either of the stimulation devices.

First-pulse-TMS was applied as previously^19^ using a conventional TMS stimulator (Magstim Rapid). Continuous-TMS was applied using the xTMS device^25^. The pulses applied by the xTMS were designed to closely approximate the biphasic pulse shape applied by the Magstim Rapid. The xTMS approximates a reference pulse using pulse-width modulation (PWM) with seven voltage levels, optimised using a model-predictive control algorithm^25^. Previous computational and in-human validation studies compared the effect of PWM pulses with their conventional reference pulses and found no differences, except in motor threshold intensity^26,27^.

For both devices, pulses were applied using a 70 mm figure-of-8 Magstim coil (Magstim Co., UK).

### Phase-locked TMS

Tremor phase was estimated in real time using an accelerometer. First, the participant’s tremor was recorded in a 3-minute baseline block, allowing us to determine the mean tremor frequency and dominant tremor axis (x, y, or z) for each participant for each session. To estimate the phase angle of the tremor for phased-lock TMS, the accelerometer data from this dominant axis were filtered with a 1Hz high-pass filter to remove orientation bias and then a phase-tracking algorithm (Oscilltrack) was applied via a microcontroller-based device, as previously described^10,22^. Having estimated tremor phase, TMS was triggered at predetermined phase angles via either the First-pulse-TMS and continuous-TMS systems.

TMS data were acquired at each of nine distinct stimulation phase bins ranging from 40° to 360°. Each of the 16 TMS blocks consisted of four TMS trains, each of which lasted approximately 3 seconds. Each train was separated by a 7 second inter-train interval. The phase for each stimulation train was randomly assigned to avoid any sequential bias that could potentially influence tremor patterns. Each of the nine phase conditions was pseudorandomly repeated approximately seven times across 16 blocks. Participants were offered a break between each block.

For First-pulse-TMS, the first pulse of the train was triggered at a specific phase of the oscillation using the OscillTrack algorithm. All subsequent pulses were then delivered at the participant’s baseline tremor frequency. Twelve TMS pulses were delivered in each train. Conversely, continuous-TMS allows continuous monitoring of tremor amplitude and frequency, enabling precise phase-locking of every pulse. This resulted in a variable number of pulses for each train, depending on changes in the frequency of the tremor.

### Data analysis

#### Determining the dominant tremor axis

Accelerometer data was sampled at 1000Hz and the channel with the maximum peak power between 4Hz-10Hz in the power spectral density (PSD) across the whole recording was used to determine the effects of TMS on tremor amplitude.

#### Post-hoc analysis of phase

We calculated post-hoc phase to determine the accuracy with which phase-locked stimulation was delivered. Accelerometer data was filtered between ± 2Hz of the peak tremor frequency in the PSD (see above) using a 2^nd^ order zero-phase lag Butterworth filter. The Hilbert transform was then used to estimate the phase of the filtered signal at each timepoint, allowing us to determine the phase at which TMS pulses had been delivered.

#### Relationship between predetermined and effective target phase

To analyse the accuracy and amplitude effects of phase-locked stimulation, it is crucial to have an accurate measurement of the phase at which stimulation was delivered (“delivered phase”). We set the closed-loop system to trigger stimulation upon detection of on one of nine *predetermined* target phases. Based on our previous studies, we did not expect any delay between detection of the predetermined target phase and stimulation^10,13^, however we observed consistent, fixed offsets between the predetermined target phase and the “effective” phase at which stimulation was delivered. These offsets were completely consistent within, but variable between, each subject and were likely due to a systematic phase lag introduced by hardware high-pass filtering (see above) and/or the selection of a different dominant tremor axis between the online tracking and the post-hoc analysis. As the difference between predetermined target phase and effective phase was consistent across *all* phases *within* subject, the effective target phase could be calculated by applying that offset to all predetermined target phases (Fig. 1). The effective target phases were then used for all analysis.

**Figure 1.**
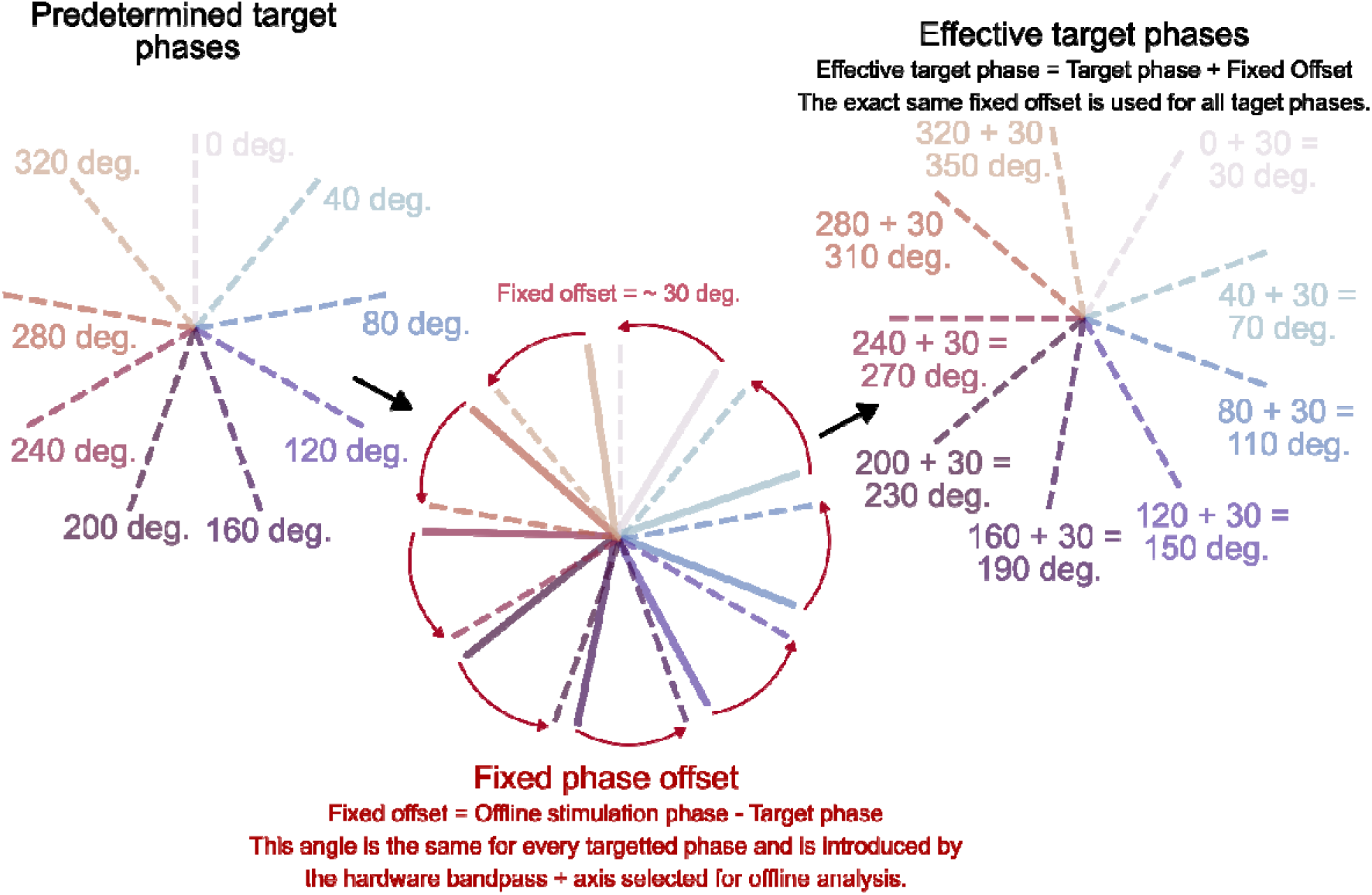
The effective target phase of stimulation accounted for consistent phase offsets within subjects. There was a systematic difference between the OscillTrack target and post-hoc analytical phase estimate of stimulation, most likely introduced by hardware high pass filtering prior to the application of the OscillTrack algorithm and differences in the selected tremor axis. To account for this, we shifted the target phases by a single, fixed angle to align them with the post-hoc determined phase. Importantly, the same fixed offset was added to all of the target phases. This has the effect of rotating all of the target phases around the circle by the same amount relative to one and other. Statistical approaches were selected such that this rotation would not affect the outcome of analysis. Using this adjustment ensures that the effects on the change in tremor amplitude are displayed in association with the phase that was being *effectively* targeted by the OscillTrack algorithm.

#### Phase precision

To evaluate the effectiveness of the OscillTrack algorithm for the online tracking of tremor phase, we determined the precision of the phase-locking of stimulation by calculating the angular error between each delivered stimulation phase and the effective target-phase. This was calculated as

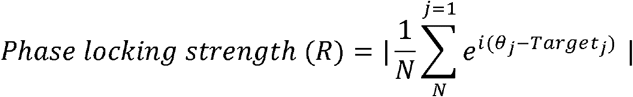

where *N* is the total number of stimulations and *θ*_*j*_ is the phase of the *j*th stimulation pulse as determined in the post-hoc analysis and *Target*_*j*_ was the adjusted target phase of the *j*th stimulation pulse. The value of R ranges from 0 (indicating stimulation was scattered randomly around the cycle) to 1 (indicating all stimulation pulses were delivered at the exact same phase). This phase-locking strength is independent of whether the predetermined or effective target-phase is used.

#### The consistency of the phase of successive stimulation pulses

To measure phase-consistency across the stimulation train, we separated TMS pulses by their target-phase and by the pulse number within the stimulation train (for the 1^st^, 2^nd^, 3^rd^, 4^th^ and 5^th^ pulse). We then calculated the phase locking strength for each pulse number separately and averaged this across the different effective target-phases. The resulting phase-locking value reflects how consistent the phase of stimulation was for each pulse of the stimulation train.

This was calculated using the mean resultant vector length, a measure of circular concentration. The formula is

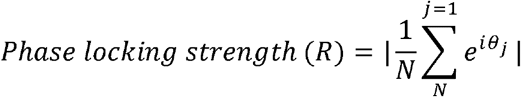

where *N* is the total number of stimulation trains at a certain target phase and *θ*_*j*_ is the phase of a set pulse number (i.e., the 1^st^, 2^nd^, 3^rd^, 4^th^ and 5^th^ pulse) in the *j*th stimulation train as determined in the post-hoc analysis. The value of R ranges from 0 (indicating stimulation was scattered randomly around the cycle) to 1 (indicating all stimulation pulses were delivered at the exact same phase). This allowed us to determine how consistent the phase of stimulation was across successive pulses of the stimulation train.

#### Target-phase dependent changes in tremor amplitude

To analyse the effect of stimulation phase on tremor amplitude, accelerometer data was band-passed filtered at ± 2Hz of peak tremor frequency in the PSD. The instantaneous amplitude of the filtered signal was then computed using the Hilbert transform, smoothed using a gaussian kernel (std=150ms, length=1s) and z-scored. For each block of stimulation, we calculated the median tremor amplitude in the 1 second preceding stimulation and the 3s blocks during which phase-locked stimulation was delivered. The change in tremor amplitude was then calculated as the difference between these two medians.

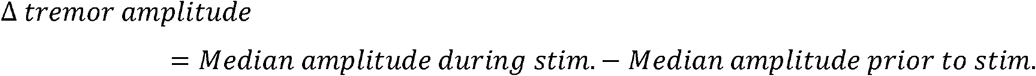

To evaluate the effect of the phase of stimulation on the change in tremor amplitude, circular-linear models were then fit to the change in tremor power as a function of the adjusted target-phase of stimulation on a patient-by-patient basis. Standard linear regression is unsuitable for modelling the effect of a circular variable, such as stimulation target-phase, on a linear outcome like tremor amplitude. A model of the form *y* = *mx* + *c* fails because it cannot account for the periodic nature of a circle, where 359° is adjacent to 0°.

To correctly model this relationship, we decompose the circular predictor into two linear components: its cosine and sine. This transforms the model into:

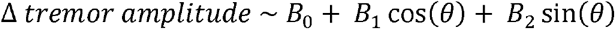

where *θ* is the stimulation target phase of each block of stimulation.

This specification is powerful because it embeds the cyclical nature of the phase directly into the regression framework. The cosine and sine terms work together to represent each phase’s unique position on a circle, correctly capturing the proximity of 0° and 359°. This allows the model to fit a smooth, periodic curve that can identify the precise phase at which the stimulation has its peak effect on tremor. This sinusoidally models the change in tremor amplitude as a function of the phase of stimulation. The magnitude and phase relationship between the change in tremor amplitude and phase of stimulation can then be determined by considering the relative values of the coefficients *B*_1_ and *B*_2_.

That is:

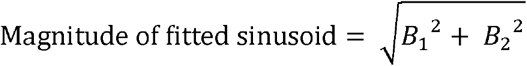

And

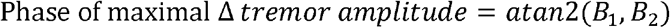

where *B*_1_ and *B*_2_ are the values of coefficient for the cos (*θ*) and sin (*θ*) terms in the model respectively (see above).

We extended these models to account for other possible confounding variables which may explain variance in the change in tremor amplitude. To control for the effect of greater cumulative number of stimulations with passing time, we included the trial number as a regressor. We also reasoned that the tremor amplitude prior to stimulation may be an important predictor of what happens in the following stimulation block. We thus include a term for the baseline amplitude of the tremor (i.e., the amplitude of the tremor in the 1 second prior to stimulation) in the model. All analyses were therefore performed using the full model:

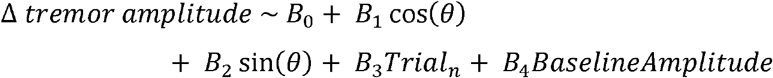

This model was fitted on a subject-by-subject basis. For each patient, this model was then compared with a null model that did not include the phase of stimulation. That is,

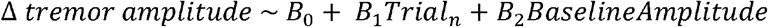

The added variance explained by including the phase of stimulation in the overall model was captured by the coefficient of partial determination (CPD):

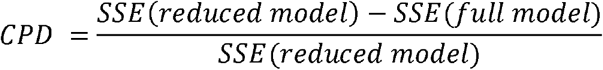

where SSE is the sum of squared errors.

To assess whether CPD was at above chance levels, we generated a null distribution by randomly shuffling the target-phase labels associated with each STIM-ON period. Circular-linear models were then fit between the change in tremor power and the shuffled phase labels:

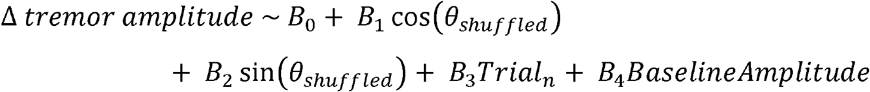

The CPD of the model using the shuffled phase-labels was then calculated as above for each patient, and this was repeated 1000 times. A null distribution for CPD pooling across patients was computed by averaging the CPD generated for each patient under null conditions (with target-phase labels shuffled). If the real patient-averaged CPD was greater than the 95^th^ percentile of the null distribution it was classified as significant. Again, the predetermined and the effective target-phase in the regression models result in an identical Coefficient of Partial Determination (CPD).

To assess the statistical significance of the confounding variables (i.e., trial number and baseline amplitude), the same procedure as above was applied separately. This was then compared to a null distribution produced by randomly shuffling the value of the confounding variable across trials and using that to generate a distribution for CPD under null conditions. If the real patient-averaged CPD was greater than the 95^th^ percentile of the patient-averaged CPD under null conditions, it was classified as significant.

## Results

We hypothesised that continuous phase-locked TMS would modulate tremor amplitude in individuals with ET. To test this, we compared the capability of cycle-by-cycle phase-locked stimulation (continuous-TMS) and first pulse phase-locked stimulation (First-pulse -TMS) to modulate the amplitude of ongoing tremor. Stimulation was delivered in alternating periods of 3s of stimulation followed by 7s of rest (Fig. 2B).

**Figure 2.**
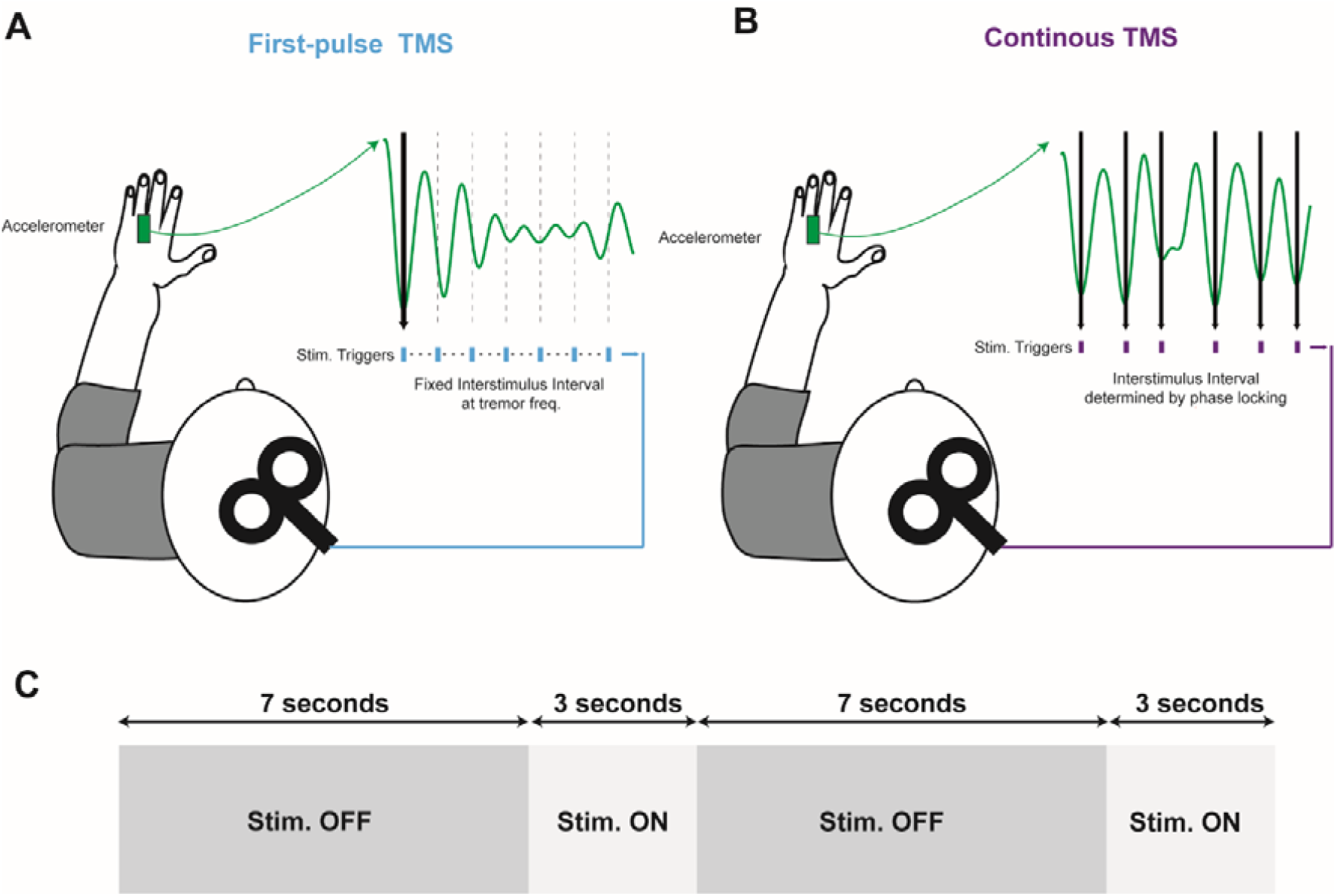
Schematic description of closed-loop stimulation protocols. Tremor was recorded with an accelerometer mounted to the dorsum of the hand and the phase of tremor was continuously tracked using the Oscilltrack algorithm, so that the tremor phase was always available. **A, B)** Stimulation was delivered using either the First-pulse-TMS or Continuous-TMS protocol. **A)** For First-pulse-TMS, the first stimulation pulse was triggered by the detection of the target phase, but the following pulses were given at fixed-intervals determined by the tremor frequency. Phase-locking of the stimulus to ongoing tremor was, therefore, dependent on matching between the frequency of the tremor and stimulus train. **B)** For Continuous-TMS, the timing of every pulse in the stimulation train is triggered by detection of the target phase in the accelerometer signal. This will result in a less regular interstimulus interval, but continuous locking of the TMS pulses to the ongoing tremor phase. **C**) For both TMS paradigms, stimulation was delivered over a 3s period, which was followed by 7s with the stimulation off.

### Continuous-TMS, but not First-pulse-TMS, delivers reliable phase-locked stimulation

We first wanted to verify that the phase-locking achieved with continuous-TMS was superior to that achieved with conventional, First-pulse-TMS, where only the first pulse of the stimulation train is truly phase-targeted (Fig. 3). We, therefore, examined the consistency of the tremor phase at which TMS pulses were delivered for each consecutive pulse in the stimulation train. As expected, continuous-TMS was consistently delivered at the effective target phase, whereas the phase of First-pulse-TMS pulses became increasingly inconsistent (Fig. 3e).

**Figure 3.**
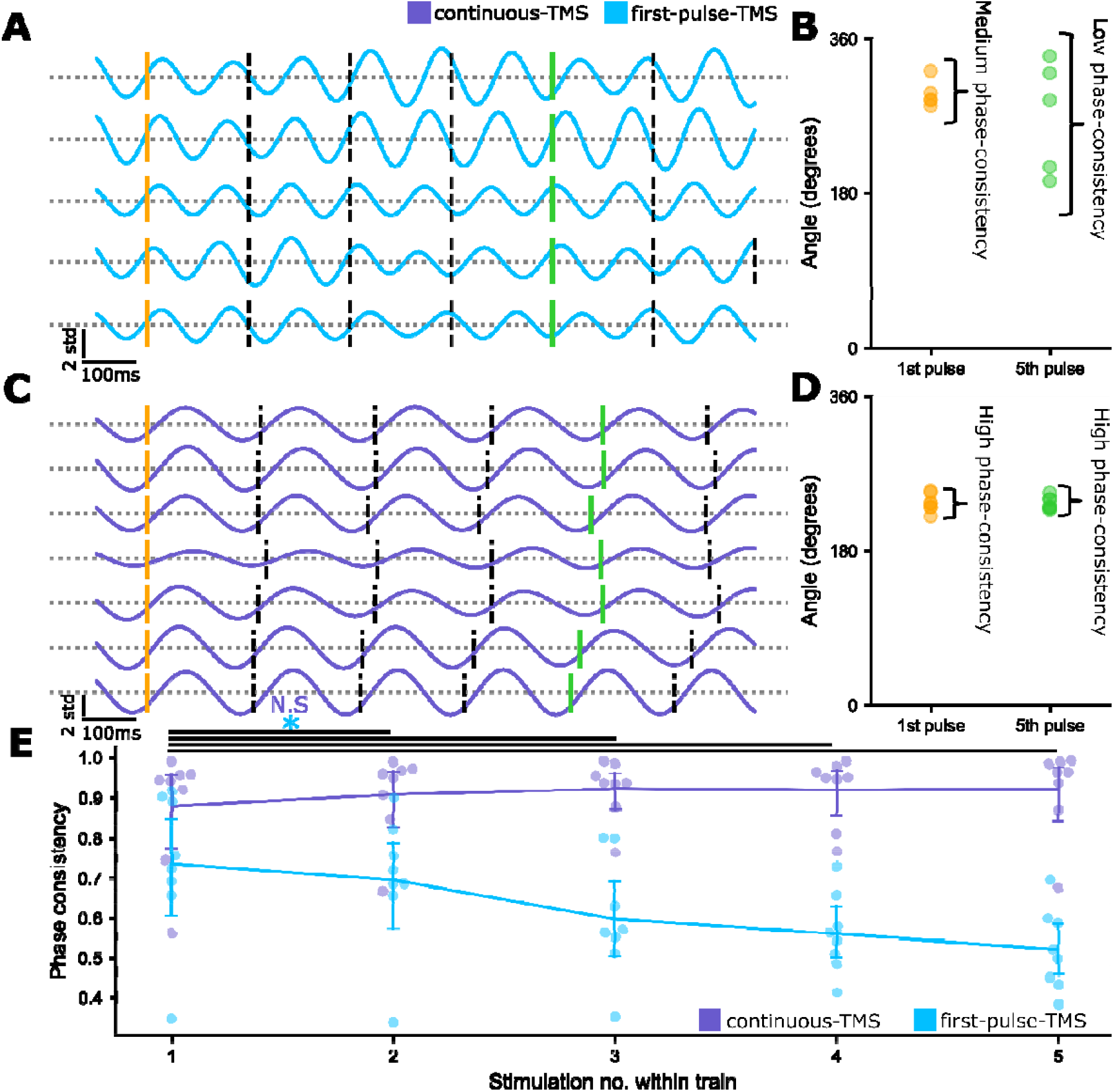
Continuous-TMS supports continuous tremor phase-targeted stimulation. **A)** Example tremor-filtered accelerometer data from all the STIM-ON periods with an adjusted target phase of ∼262 degrees (internal target = 280 degrees) in a single First-pulse -TMS session. In this session, there were four such STIM-ON periods. Stimulation timings are marked with vertical lines. The First-pulse stimulation pulse was marked with an orange line, whereas the 5^th^ was marked with a green line. **B)** A scatter plot of the phase of the First-pulse (orange) and 5^th^ (green) First-pulse -TMS pulse in the example in **A**. A moderate degree of consistency in the phase of stimulation is found for the First-pulse pulse of the stimulation train, with a much lower phase consistency by the 5^th^ pulse. **C)** Example tremor-filtered accelerometer data from all of the STIM-ON periods with an adjusted target phase of ∼269 degrees (internal target = 320 degrees) in a single Continuous-TMS session. In this session, there were 7 such STIM-ON periods. Stimulation timings are marked with vertical lines. The First-pulse stimulation pulse was marked with an orange line, whereas the 5^th^ was marked with a green line. **D)** A scatter plot of the phase of the First-pulse (orange) and 5^th^ (green) Continuous-TMS pulse in the example in **C**. A high degree of consistency in the phase of stimulation is found for the First-pulse and 5^th^ pulse of the example stimulation trains. **E)** Phase-consistency remains stable across the stimulation train with Continuous-TMS (First-pulse vs. 2^nd^, 3^rd^, 4^th^ and 5^th^ pulse of the stimulation train, p>0.05 Wilcoxon signed-rank test, multiple comparison correction using Benjamini-Hochberg), but degrades with increasing stimulations in First-pulse -TMS (First-pulse vs. 2^nd^, 3^rd^, 4^th^ and 5^th^ pulse of the stimulation train, p<0.05 Wilcoxon signed-rank test, multiple comparison correction using Benjamini-Hochberg). A post-hoc calculation of phase using the Hilbert transform is used for all of the above analysis.

We next quantified how these differences translated into *overall* phase-locking accuracy. In each participant, continuous-TMS, but not First-pulse-TMS, pulses were delivered accurately across all 9 effective target phases (Fig. 4a and 4b). When all phases were pooled by calculating the error in relation to the target phase, it was evident that phase-locking accuracy was considerably greater during continuous-TMS than First-pulse-TMS (Fig. 4c and 4d). Indeed, the mean phase locking (i.e. consistency of phase at which stimulation was delivered) was significantly greater during continuous-TMS than First-pulse-TMS (Fig. 4e, Mann-Whitney U-test, p<0.001).

**Figure 4.**
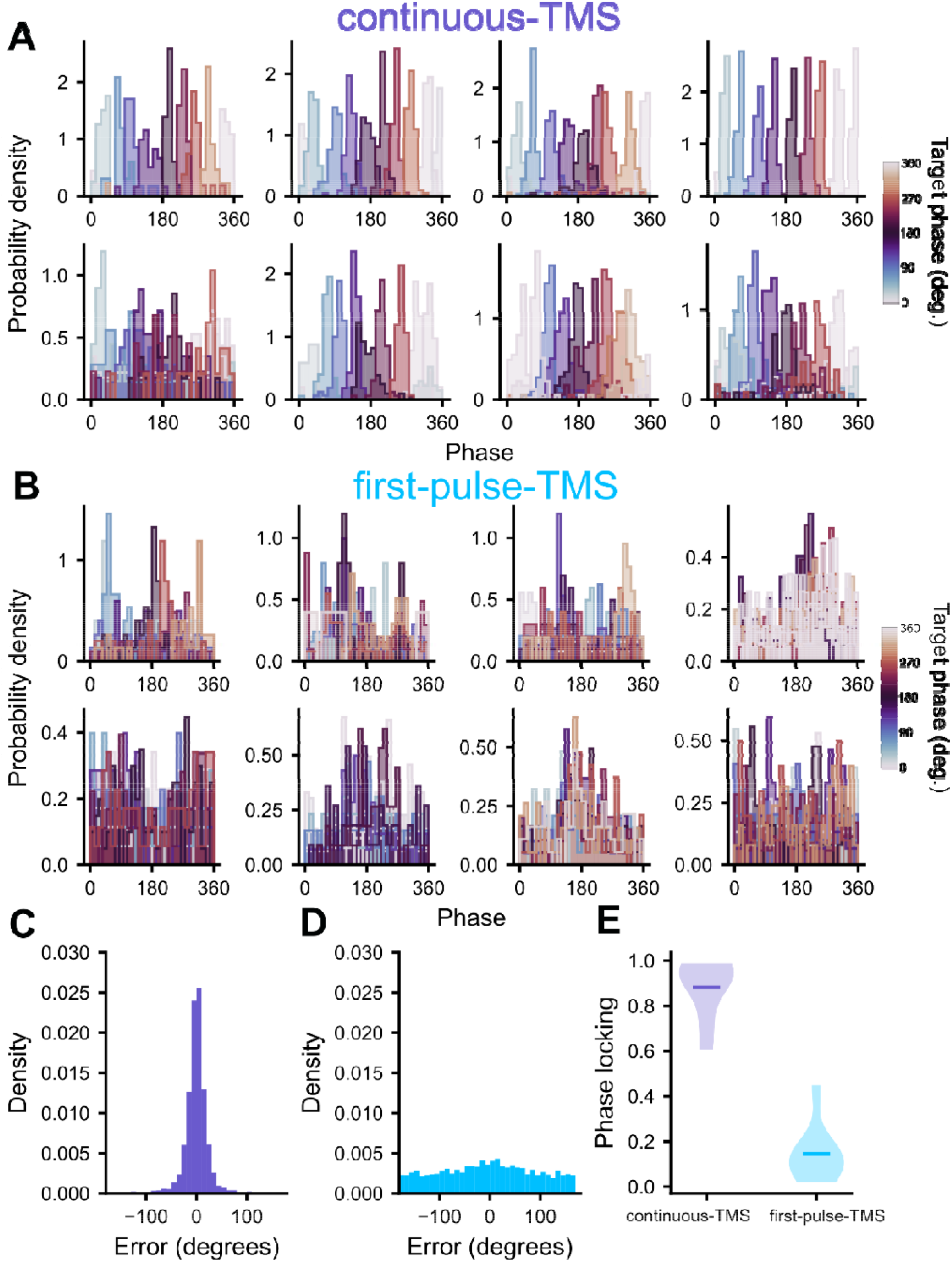
Accurate phase-locking to tremor oscillations could be achieved with Continuous-TMS but not First-pulse -TMS. **A)** Histograms for each patients displaying the phase of the tremor oscillations (calculated post-hoc) that each of the *Continuous-TMS* pulses were delivered at. Different coloured bars represent the different target phase conditions recorded in patients. **B)** Histograms for each patients displaying the phase of the tremor oscillations (calculated post-hoc) that each of the *First-pulse -TMS* pulses were delivered at. Different coloured bars represent the different target phase conditions recorded in patients. **C)** The error between the adjusted target-phase and the post-hoc calculated phase of *Continuous-TMS* pooling all stimulations across the 8 patients. **D)** The error between the adjusted target-phase and the post-hoc calculated phase of *First-pulse -TMS* pooling all stimulations across the 8 patients. **E)** Violin plot showing the strength of phase locking of stimulation to tremor in Continuous-TMS and First-pulse -TMS, with the mean phase locking coefficient averaging across all patients being represented by a horizontal line. The strength of phase locking was calculated individually for each of the patients (n=8 Continuous-TMS, n=8 First-pulse -TMS). When accurate phase locking is achieved, angular error should be tightly distributed around 0 phase (as in **C**). Weaker phase locking results in a more uniformly distributed angular error (as in **D**). The circular concentration of the phase of stimulation was significantly greater in Continuous-TMS than First-pulse -TMS (Mann-Whitney U-test, p<0.001).

### Continuous-TMS, but not First-pulse-TMS, resulted in bidirectional modulation of tremor amplitude

Next, we investigated whether the effect of stimulation on tremor amplitude was dependent on the target phase. To do this, we fitted circular-linear models for each patient to determine the relationship between target-phase and change in tremor amplitude. To determine if the phase of stimulation had a statistically significant effect, we calculated the *additional* variance explained by including target-phase in the model compared with a null distribution generated by repeatedly fitting circular-linear models with shuffled phase labels. We included baseline tremor amplitude and the trial number as covariates in our regression models to control for potential confounding effects.

Continuous-TMS sinusoidally modulated tremor amplitude as a function of target phase for several of the patients (Fig. 5a). Including the sinusoidal effect of target-phase on change in tremor amplitude significantly increased model fits (Fig. 5b, permutation test, p=0.0024), demonstrating that the target-phase of stimulation sinusoidally modulated the phase of tremor amplitude. Including the baseline amplitude also explained significantly more variance than the null distribution, suggesting that a “return to the mean” accounts for some of the amplitude modulation (Fig. 5B, permutation test, p<0.001). Importantly, however, our approach controls for this effect on tremor amplitude when considering target phase. Including this confounder within circular-linear models therefore allowed us to separate the effects of the phase of stimulation from the tendency of tremor amplitude to return from high or low values towards the mean. Finally, trial number (i.e. the order in which the target phases were applied) did not significantly explain amplitude modulation, suggesting that effects such as fatigue were not a major factor (Fig. 5B, permutation test, p=0.15).

**Figure 5.**
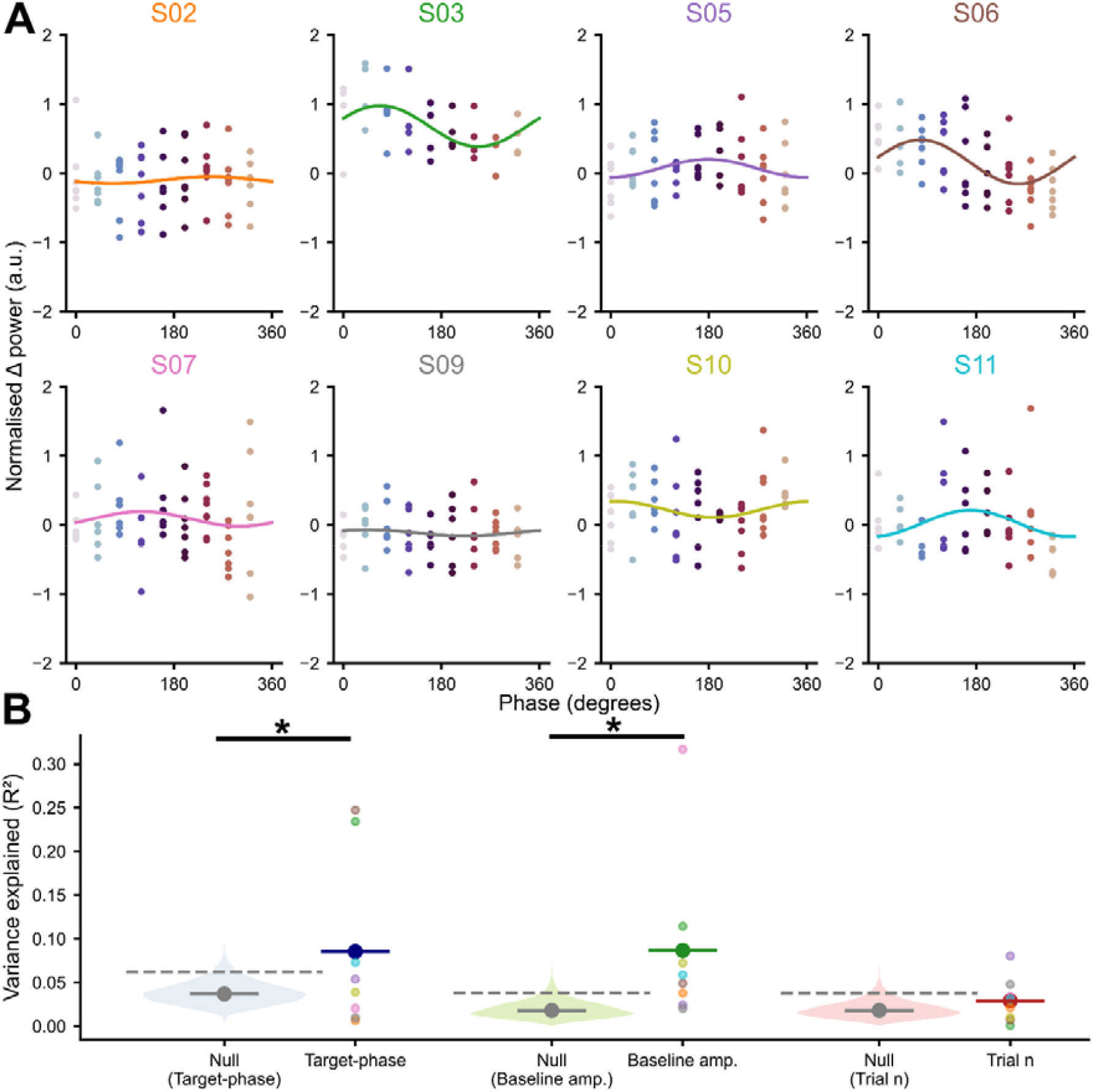
Phase-locked Continuous-TMS can bidirectionally modulate the amplitude of tremor: **A)** The change in tremor amplitude as a function of the targeted phase of the Continuous-TMS stimulation for each ET patient (n=8). The change in tremor was calculated as the difference between the tremor amplitude in the 3s STIM-ON period and the preceding 1s of STIM-OFF. Each point here represents a single STIM-ON block. A circular-linear model was fitted between the targeted phase of Continuous-TMS and the change in tremor amplitude. Variance accounted for by confounding variables (i.e., trial n and baseline amplitude) was subtracted from each change in amplitude scatter point for plotting. The sinusoid fit to the change in tremor power in the circular linear model is displayed as a solid line. **B)** The additional variance in the change in tremor amplitude added by including each independent variable in the circular-linear models (CPD). Left to right, this is the target phase of stimulation, the baseline amplitude of the tremor and the trial number. For each independent variable, on the right the CPD for each patient was plotted as a scatter point with a colour that corresponds to **A** and the overall mean of the CPD pooling across patients was represented by a horizontal line. To the left of this, for each independent variable, was a null distribution for the patient pooled mean CPD. The median value of this null distribution was marked with a gray filled horizontal bar and the 95^th^ percentile was marked with a gray dashed line. The CPD for target-phase (p=0.0024) and baseline amplitude (p<0.001) was significantly greater than would be expected under null conditions. The CPD for trial number was not statistically significant (p=0.15).

Unlike continuous-TMS, no participant exhibited a systematic relationship between the target phase and the change in tremor amplitude in response to First-pulse-TMS (Fig. 6a); the added variance in tremor amplitude explained by stimulation phase was well below the 95^th^ percentile of the null distribution (Fig. 6b, permutation test, p=0.94). In contrast, both trial number (permutation test, p=0.0086) and baseline amplitude (permutation test, p<0.001) significantly explained variance in tremor amplitude compared to the null distribution (Fig. 6B). In the case of baseline amplitude, this indicates that “regression to the mean” effects were common to both TMS protocols and were therefore a general feature of the experimental paradigm. This further highlights the importance of controlling for these factors within the models, allowing us to control for the First-pulse tremor amplitude and any slow drifts in tremor power that occur over the recording. Overall, these results suggest that continuously phase-targeted stimulation was necessary to modulate tremor amplitude in a phase-dependant manner.

**Figure 6.**
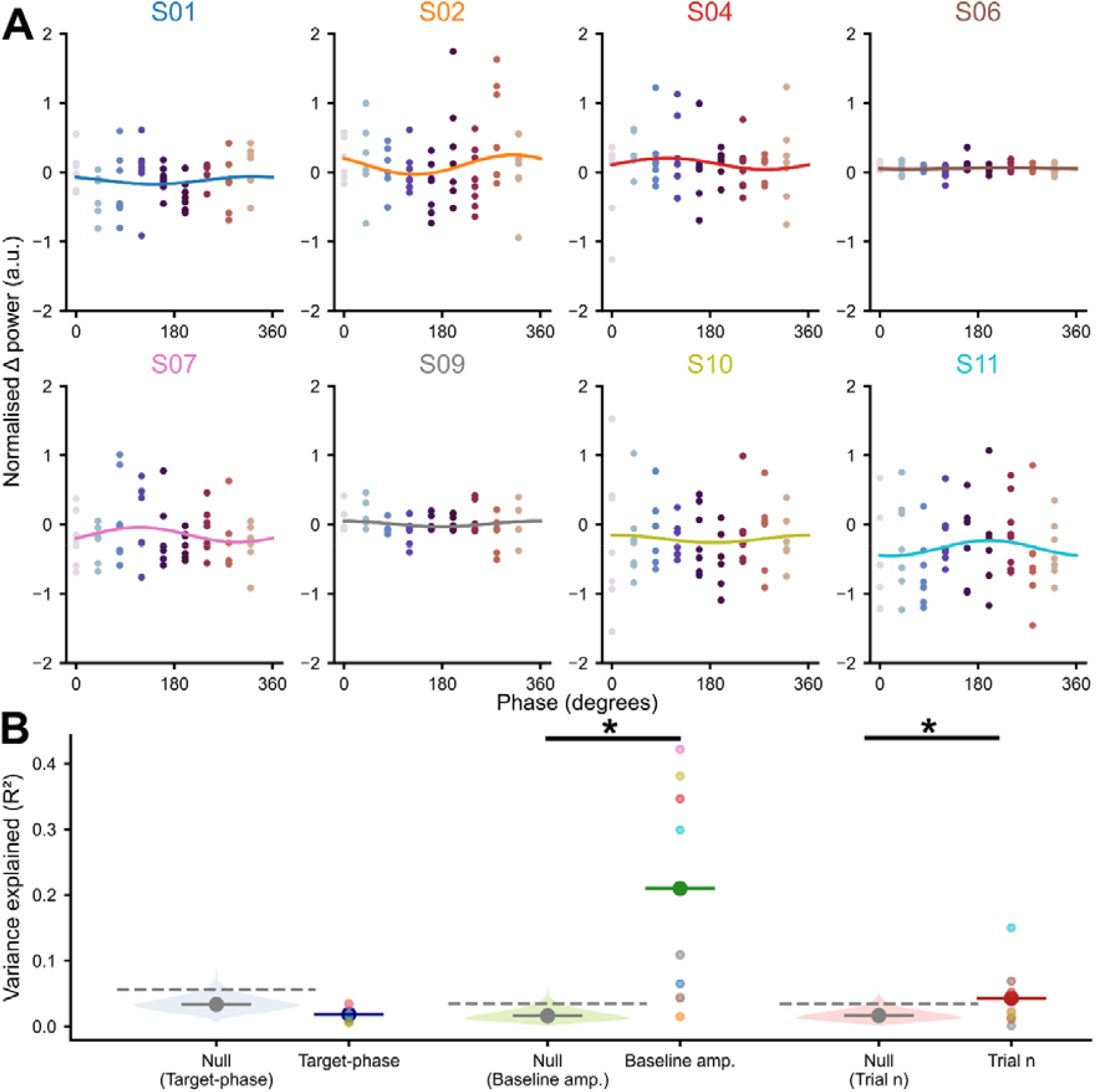
First-pulse-TMS did not lead to phase dependent modulation of tremor amplitude: The change in tremor amplitude as a function of the targeted phase of the First-pulse -TMS stimulation for each ET patient (n=8). The change in tremor was calculated as the difference between the tremor amplitude in the 3s STIM-ON period and the preceding 1s of STIM-OFF. Each point here represents a single STIM-ON block. A circular-linear model was fitted between the targeted phase of First-pulse -TMS and the change in tremor amplitude. Variance accounted for by confounding variables (i.e., trial n and baseline amplitude) was subtracted from each change in amplitude scatter point for plotting. The sinusoid fit to the change in tremor power in the circular linear model is displayed as a solid line. **B)** The additional variance in the change in tremor amplitude added by including each independent variable in the circular-linear models (CPD). Left to right, this is the target phase of stimulation, the baseline amplitude of the tremor and the trial number. For each independent variable, on the right the CPD for each patient was plotted as a scatter point with a colour that corresponds to **A** and the overall mean of the CPD pooling across patients was represented by a horizontal line. To the left of this, for each independent variable, was a null distribution for the patient pooled mean CPD. The median value of this null distribution was marked with a gray filled horizontal line and the 95^th^ percentile was marked with a gray dashed line. The CPD for baseline amplitude (p<0.001) and trial number (p=0.0086) was significantly greater than would be expected under null conditions. The target-phase of stimulation did not significantly affect the tremor amplitude here (p=0.94).

### Phase-locked TMS leads to long-term tremor suppression in some participants

This study was designed to investigate the immediate effects of phase-locked TMS on tremor amplitude, rather than to test long-lasting effects of distinct stimulation trains. As an exploratory analysis, we examined potential long-lasting effects of stimulation trains in a subset of participants showing a marked tremor reduction during the experiment. In these participants, a prolonged suppression of tremor outlasting the stimulation itself could be achieved, both with continuous-TMS (Fig 7a, b) and First-pulse-TMS (Fig 7c, d). We examined periods of time where participants had returned to the stimulation protocol after a break of at least 20s. On returning to the stimulation protocol, these participants exhibited a profound reduction in tremor amplitude that lasted well into subsequent STIM-OFF and STIM-ON periods, highlighting possible plastic changes in the tremor generating circuits. Notably, in one participant, a reduction in tremor amplitude of ∼80% was achieved over the course of around 30s of the stimulation protocol (Fig 7c, d, supplementary video 1). The effects observed in these participants highlight that phase-locked TMS can achieve tremor suppression lasting beyond immediate stimulation, potentially leading to clinically meaningful improvements in tremor severity.

**Figure 7.**
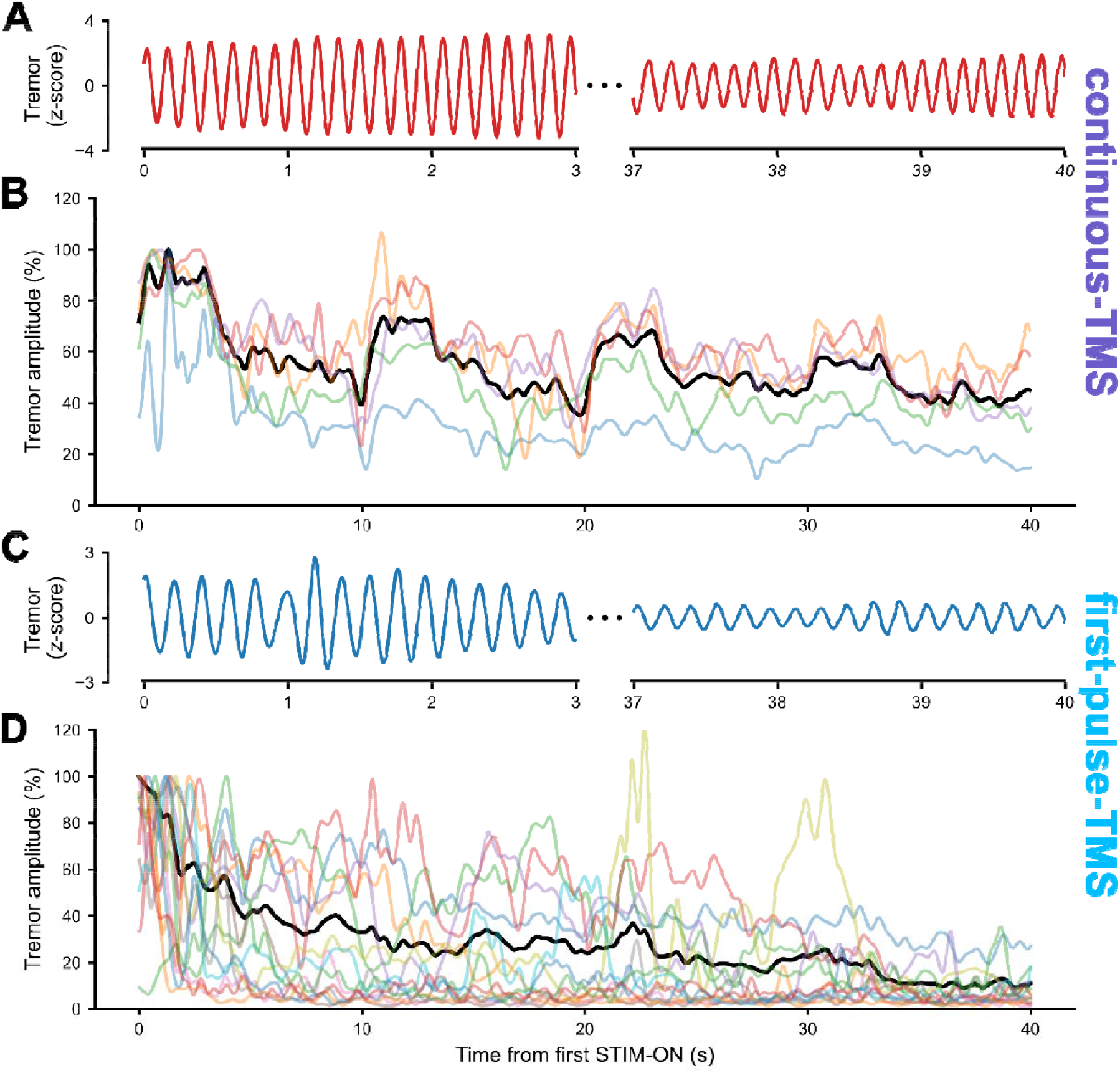
Both continuous-TMS and first-pulse-TMS were associated with long lasting suppression in a subset of participants: Here we show the effects of TMS over longer time frames from 2 example participants, one recorded in each of continuous-TMS and first-pulse-TMS. We analyse periods of time where participants recommence the stimulation protocol after a break of at least 20s. Tremor amplitude was evaluated across the 40s from the delivery of stimulation. This 40s would span 4 distinct STIM-ON and STIM-OFF periods (see Fig 2A). **A)** A representative example of tremor during the 0-3s and 37-40s after subject S04 recommenced the continuous-TMS stimulation protocol following a break (colour matched to the corresponding tremor amplitude in **B**). **B**) The tremor amplitude during all of the occasions that participant S04 recommenced the continuous-TMS stimulation protocol (coloured) and the mean of these events (black). Both were expressed as a percentage of the maximal tremor amplitude in the first 5s of recommencing stimulation. **C)** A representative example of tremor during the 0-3s and 37-40s after subject S03 recommenced the first-pulse-TMS stimulation protocol following a break (colour matched to the corresponding tremor amplitude in **D**). **D**) The tremor amplitude during all of the occasions that participant S03 recommenced the first-pulse-TMS stimulation protocol (coloured) and the mean of these events (black). Both were expressed as a percentage of the maximal tremor amplitude in the first 5s of recommencing stimulation.

## Discussion

Phase-targeted stimulation has considerable potential to harness temporal dynamics for acute and chronic modulation of neural oscillations and the underlying pathological manifestations such as tremor. Here we demonstrate in individuals with ET that continuous-TMS using the xTMS device allows the delivery of accurate, continuous phase-targeted stimulation, and resulted in a phase-dependent modulation of tremor amplitude that could not be achieved with First-pulse-TMS. These results highlight the potential of phase-targeted TMS to modulate disease-related activity in ET and other brain disorders and to reveal pathophysiological mechanisms and develop novel therapies.

### Phase-targeted TMS can bidirectionally modulate the amplitude of tremor in ET

The central finding of this study is that phase-targeted TMS can directionally modulate the tremor amplitude in ET. This adds to two previous studies^7,21^ demonstrating that phase-targeted deep brain stimulation and transcranial alternating current stimulation (tACS) achieved the same effect. In the study by Brittain and colleagues, tACS aligned to the suppressing phase could achieve a mean 50% reduction in tremor amplitude when stimulation was applied for 30s. Similarly, phase-targeted DBS outperforms high frequency stimulation^21^.

Here, we set out to define differential phase-effects on tremor amplitude across the cycle, meaning that our levels of modulation were more modest. Although 3 second stimulation blocks inevitably led to more noisy data, it allowed us to analyse the consistency of our effects across multiple stimulation blocks for each target phase and to define amplitude changes against baseline tremor immediately before stimulation. In conjunction with circular/linear modelling, this allowed detailed characterisation of the amplitude response. As in our previous studies of parkinsonian beta oscillations^10^ and hippocampal theta oscillations^13^, this modulation was broadly sinusoidal with the amplifying and suppressing phases being roughly half-a-cycle apart.

A general confound of phase-targeted stimulation is that, by locking stimulation to consecutive cycles of the target oscillation, it will always lead to a regular stimulation pattern at the frequency of the input signal. Modulation may therefore result from the “entraining” effects of the stimulation pattern, rather than closed-loop interaction. We have previously controlled for this confound by “playing back” the stimulus train generated by closed-loop stimulation. We did not use that control condition here as the First-pulse-TMS protocol performed a similar role. Indeed, as the first few pulses of each stimulation train were consistently applied at the same target phase as the Continuous-TMS condition, whereas open-loop replay has no relation to ongoing phase, First-pulse-TMS is arguably an even more conservative control. Moreover, as the stimulation train was applied at the target frequency, the stimulation pattern of the First-pulse -TMS was more regular than the Continuous-TMS, where the phase-tracking caused the pulse timing to adapt to the phase-slips of the tremor. If our effects were dependent on “entraining” effects, we would expect to see significant effects of First-pulse -TMS on tremor amplitude, which are not observed. We can therefore conclude that amplitude modulation seen here was dependent on continuous closed-loop effects, rather than stimulation at tremor frequency.

### Cycle-by-cycle phase-locked TMS allows continuous tremor-phase targeted stimulation

Several recent studies have demonstrated the ability of continuous phase-targeted DBS and optogenetic stimuli to modulate ongoing neural oscillations^8-14,16,28-30^. There have been two main obstacles to employing these approaches using TMS. Firstly, the majority of studies have used either local field potentials (LFPs) or EEG as the input signal to the closed-loop. While EEG provides the obvious neural signal for non-invasive experiments, the artifacts and evoked potentials generated by transcranial electrical stimulation methods, including TMS, are large enough to degrade the ongoing phase-tracking for a given oscillation. As others have done^7,21^, we bypassed this issue by using a peripheral signal such as tremor as the input to the closed-loop, which is not corrupted by stimulation pulses. Second, while isolated/First-pulse TMS pulses have been phase-targeted previously, it has not been possible to deliver them to several consecutive cycles. This is an important distinction, as continuous locking is needed to hold an oscillation in an amplified or suppressed state^10,13^. By tracking the phase of the accelerometer signal in real time with the Oscilltrack algorithm and using the output to drive the xTMS device, we were able to achieve such cycle-by-cycle targeting. Together, this approach allowed us to provide the first example of TMS continuously targeted to an ongoing neural oscillation.

### Therapeutic potential of phase-targeted TMS

Tremor is arguably the purest example of an “oscillopathy,” whereby highly rhythmic interactions between a network of brain areas, in this case the cerebello-thalamo-cortical circuits, lead directly to symptoms. Here, we provide further evidence that phase-dependent manipulations provide a highly targeted method of directly ameliorating symptom generating neural activity. An important question for closed-loop stimulation is identifying the most effective node within the network to target with stimulation. While studies using tACS likely affected large areas of the motor cortex^7^ or the cerebellum^8^, TMS allows more focussed targeting, allowing us to conclude that primary motor cortex is a viable node at which to intervene in ET in future studies and trials.

Previous open-loop TMS studies targeting the primary motor cortex have already demonstrated a modest reduction in tremor amplitude in individuals with ET^31^. However, such stimulation was delivered independently of the tremor’s oscillatory phase or the underlying neural activity, potentially resulting in pulses administered during both tremor-amplifying and tremor-suppressing phases. In the present study, we demonstrate that our continuous-TMS approach allows reliable, phase-specific stimulation of the tremor cycle, selectively engaging distinct phases associated with either amplification or suppression of tremor amplitude. This approach enables the empirical identification of the phase at which stimulation produces maximal tremor suppression for each individual, thereby supporting the development of personalized, phase-specific TMS protocols to optimize clinical efficacy in neuromodulation.

However, for therapeutic relevance, stimulation would need to produce tremor suppression that is sustained beyond the immediate period of delivery. Our preliminary proof-of-concept analysis suggests that repeated phase-targeted stimulation may induce cumulative suppression that persists across stimulation cycles, supporting the possibility that longer or repeated interventions could yield therapeutically meaningful effects.

Future studies using phase-targeted TMS could further address several questions to strengthen the translational and therapeutic implications. Firstly, what is the acute effect of sustained (over several minutes) phase-targeted stimulation on tremor amplitude? While acute TMS is not a viable therapeutic strategy, such experiments would provide information as to the potential of motor cortical intervention in ameliorating online tremor. Secondly, longer periods of stimulation at the optimal tremor-suppressive phase could be used to examine whether sustained phase-locking can induce plastic effects. Continuously aligning stimulation to intrinsic dynamics provides the potential to alter connectivity between network nodes^33^. If phase-targeted stimulation weakens connectivity in the tremor network, it should manifest as suppression that significantly outlasts the period of closed-loop stimulation. Such effects would be necessary for the approach to have tractable therapeutic potential. In conclusion, although thalamic DBS provides an effective treatment for ET, the widespread use of TMS for the treatment of depression provides proof-of-principle evidence that such an approach is feasible and scalable^34^ and could be translated to tremor therapy.

## Abbreviations

aMT: active motor threshold
APB: abductor pollicis brevis
Continuous-TMS: cycle-by-cycle phase-locked TMS
CPD: coefficient of partial determination
DBS: deep brain stimulation
EMG: electromyography
ET: essential tremor
FDI: first dorsal interosseous
FFF: forearm finger flexors
FFE: forearm finger extensors
First-pulse-TMS: first-pulse phase-locked TMS
LFP: local field potential
NIBS: non-invasive brain stimulation
PSD: power spectral density
rMT: resting motor threshold
tACS: transcranial alternating current stimulation
TMS: transcranial magnetic stimulation

## Data availability

The dataset of this paper (https://doi.org/10.60964/rnd-0qp8-9458) is not yet openly available, as it is being used in ongoing projects. We welcome enquires for sharing this as part of a collaboration, please contact. Code highlighting key analysis will be made available on acceptance.

## Acknowledgements

We would like to thank the National Tremor Foundation for supporting the recruitment of participants for this study.

